# COVID-19 associated autoimmunity is a feature of severe respiratory disease - a Bayesian analysis

**DOI:** 10.1101/2021.02.17.21251953

**Authors:** Uriel Trahtemberg, Robert Rottapel, Claudia C dos Santos, Alex P Di Battista, Arthur S. Slutsky, Andrew J Baker, Marvin J Fritzler, on behalf of the COVID19 Longitudinal Biomarkers of Lung Injury (COLOBILI) study group

**Author notes:** Corresponding author: Marvin J. Fritzler PhD MD, Cumming School of Medicine, University of Calgary, 3330 Hospital Dr. NW, Calgary, AB, Canada T2N 4N1.

## Abstract

**Background:** Serological and clinical features with similarities to systemic autoimmunity have been reported in severe COVID-19, but there is a lack of studies that include contemporaneous controls who do not have COVID-19.

**Methods:** Observational cohort study of adult patients admitted to an intensive care unit with acute respiratory failure. Patients were divided into COVID^+^ and COVID^−^ based on SARS-CoV-2 PCR from nasopharyngeal swabs and/or endotracheal aspirates. No COVID-19 specific interventions were given. The primary clinical outcome was death in the ICU within 3 months; secondary outcomes included in-hospital death and disease severity measures. Measurements including autoantibodies, were done longitudinally. ANOVA and Fisher’s exact test were used with α=0.05, with a false discovery rate of q=0.05. Bayesian analysis was performed to provide credible estimates of the possible states of nature compatible with our results.

**Results:** 22 COVID^+^ and 20 COVID^−^ patients were recruited, 69% males, median age 60.5 years. Overall, 64% had anti-nuclear antibodies, 38% had antigen-specific autoantibodies, 31% had myositis related autoantibodies, and 38% had high levels of anti-cytokine autoantibodies. There were no statistically significant differences between COVID^+^ and COVID^−^ for any of the clinical or autoantibody parameters. A specific pattern of anti-nuclear antibodies was associated with worse clinical severity for both cohorts.

**Conclusions:** Severe COVID^+^ patients have similar humoral autoimmune features as comparably ill COVID^−^ patients, suggesting that autoantibodies are a feature of critical illness regardless of COVID-19 status. The clinical significance of autoimmune serology and the correlation with severity in critical illness remains to be elucidated.

## Introduction

The immune response in SARS-CoV2 infection shows evidence of immune dysregulation in patients with severe clinical manifestations^1^. While there is continued debate about the nature of the heightened inflammatory responses in severe COVID-19 patients^2, 3^ there is mounting evidence that some clinical and serological features bear remarkable similarities to systemic autoimmune conditions^4-6^. In addition, some COVID-19 patients continue to develop *de novo* clinical signs and symptoms reminiscent of autoimmune diseases during convalescence^7^. Potential mechanisms leading to autoimmunity in COVID-19 include increased release of self-antigens as a result of tissue damage, neutrophil activation and NETosis^8^, molecular mimicry from homologous sequences of SARS-COV2 with human proteins^9^ and activation of autoreactive immune cells due to cytokinemia^3^.

A significant limitation of studies that have reported the emergence of autoantibodies (AAB) in COVID-19^4, 6, 10^ has been the lack of control groups with similar clinical characteristics and the lack of longitudinal data monitoring the development of autoimmunity over time. Our study provides evidence that, when observed longitudinally, severe COVID-19 patients have similar AAB prevalence as a control cohort of critically ill patients.

## Methods

### Study design

This report is part of the COLOBILI study – Coronavirus Longitudinal Biomarkers in Lung Injury, an observational cohort study that includes analysis of biological samples conducted at St. Michael’s Hospital (Toronto, ON, Canada). The study was approved by the Research Ethics Board of St. Michael’s Hospital (REB# 20-078). Informed consent was obtained from the patients or their legal representatives. In cases where it was not possible to obtain consent, patients were enrolled using a deferred consent model and kept in the study until they regained capacity, or a surrogate decision maker was identified.

The inclusion criteria were all patients above age 18 years admitted to the Medical-Surgical or Trauma-Neuro intensive care units (ICU) with acute respiratory distress, suspected to have COVID-19. The exclusion criteria were refusal to participate, inability to ascertain mortality status during the first 2 weeks of the study, failure to obtain a blood sample on either day 0 or 1, or individuals known to have had COVID-19 in the 4 weeks prior to admission in any setting. Patients were followed for up to 3 months if they were in hospital or until hospital discharge, whichever occurred first. The primary outcome was death in the ICU; secondary outcomes included death outside the ICU, ICU utilization metrics, and organ dysfunction measures and scores (see Table 1). COVID19 status of all patients was determined according to diagnostic PCR of nasopharyngeal swabs and/or endotracheal aspirates. Further PCR tests were repeated by the clinical and infection control teams at their discretion to confirm negativity or if there was suspicion of a false negative result based on clinical observations. All patients in the PCR negative cohort had at least two negative tests performed acutely. One patient had only one test done acutely.

**Table 1:**
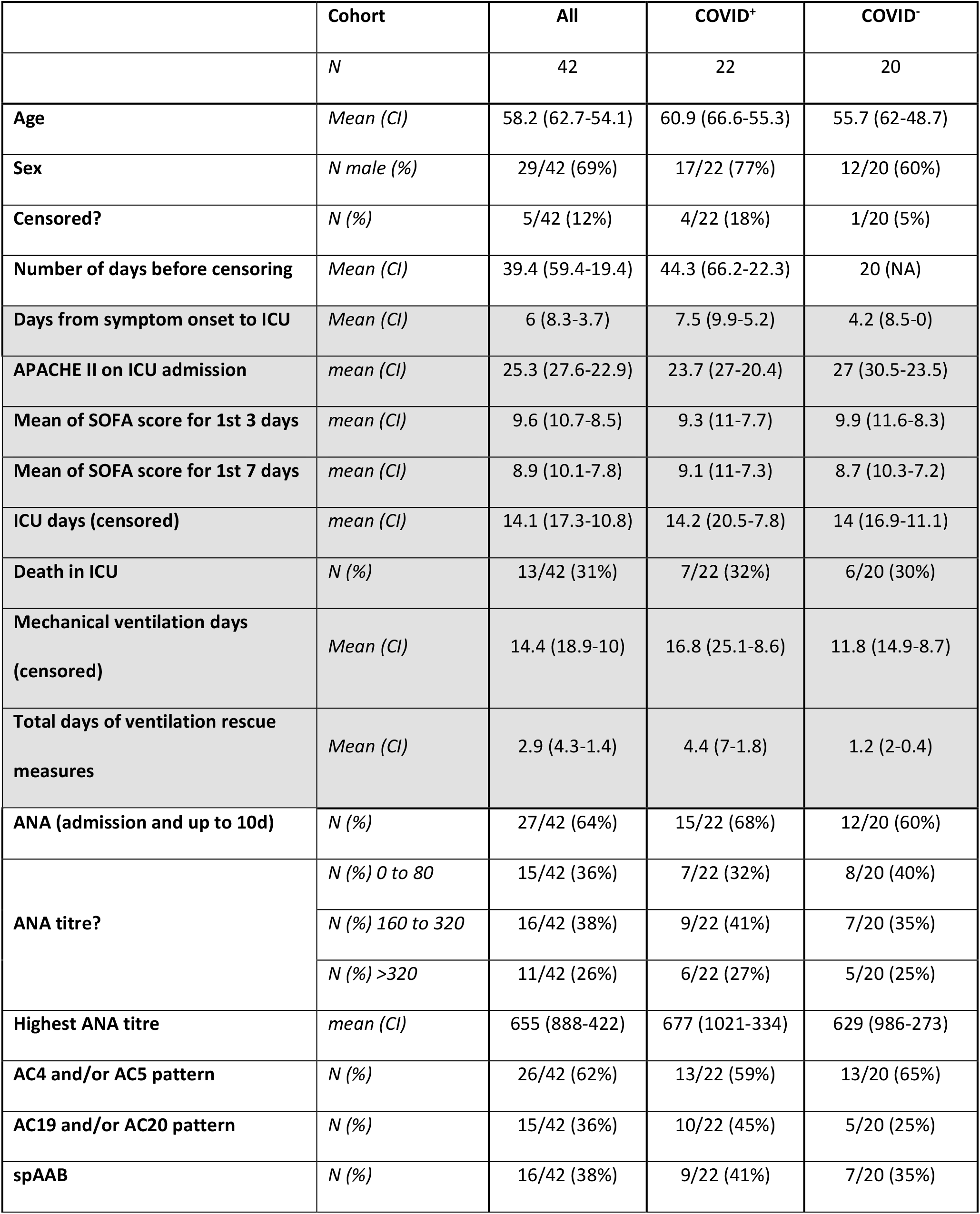
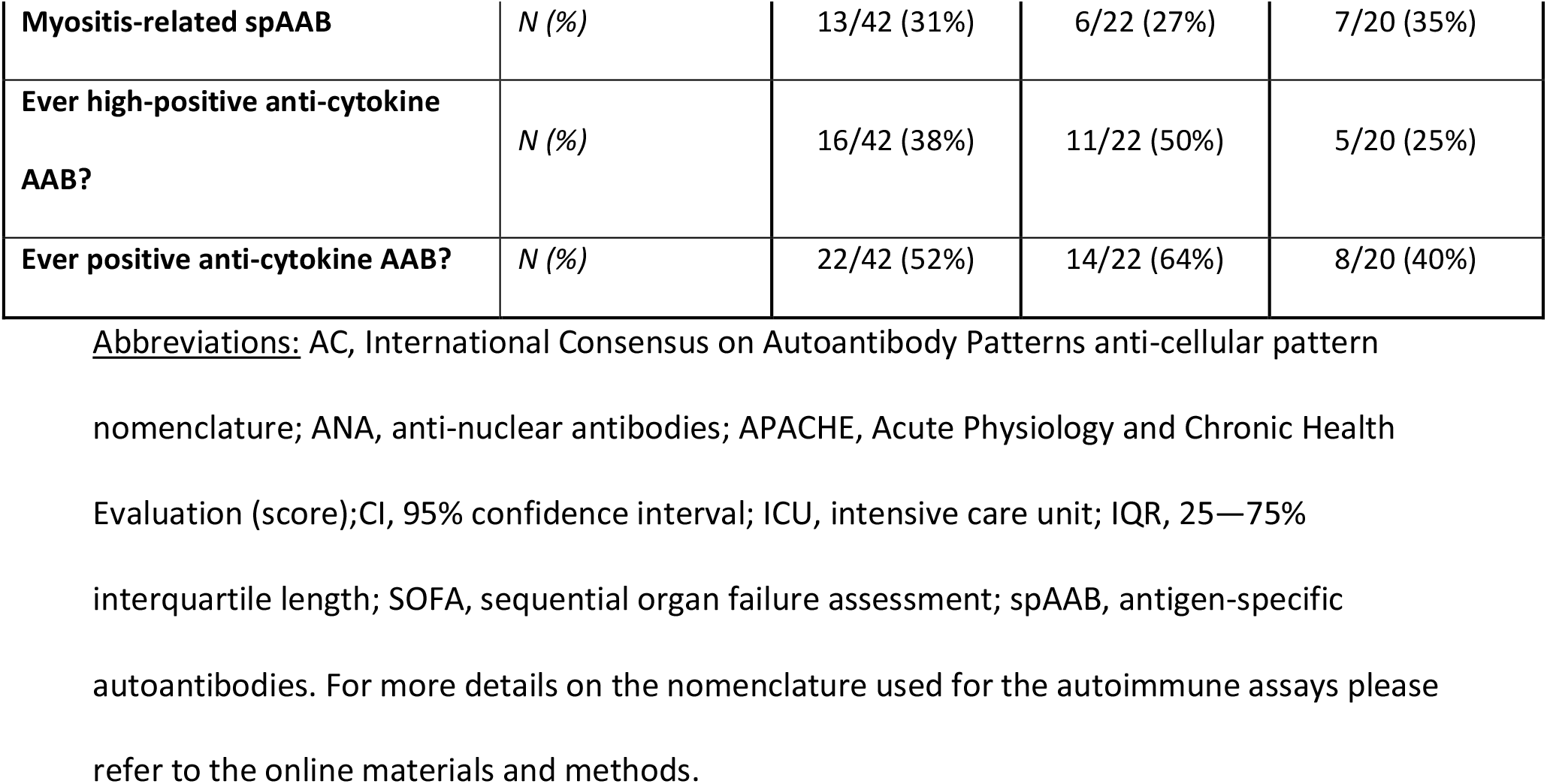
Patient Demographic, Clinical and Autoantibody Status <u>Legend:</u> The data was censored on May 31st. Days from symptom onset were self-reported by the patients or their representatives. ANA were determined with immunofluorescence using Hep-2 cells and classified using the International Consensus on Autoantibody Patterns system. The table reports the highest value found during longitudinal sampling up to day 10 of ICU admission, for standardization among patients. Titres of 0 to 80 are considered negative and titres above 320 are considered high. The SOFA score was performed daily for all patients; the average was calculated for the first 3 and 7 days in the ICU for each patient, and the mean of those averages are reported. For patients that underwent tracheostomy, mechanical ventilation days are counted until successfully weaned from ventilatory support for 24 hrs. Rescue measures included use of paralytics, proning and NO (counted additively if more than one intervention used in the same day). The clinical outcomes were measured for up to 3 months. There was no statistically significant difference between COVID^+^ and COVID^−^ patients for all variables, using ANOVA for continuous variables and Fisher’s exact test for categorical variables at α=0.05, followed by the false discovery rate at q=0.05.

|  | Cohort | All | COVID<sup>+</sup> | COVID<sup>-</sup> |
| --- | --- | --- | --- | --- |
|  | *N* | 42 | 22 | 20 |
| Age | *Mean (CI)* | 58.2 (62.7-54.1) | 60.9 (66.6-55.3) | 55.7 (62-48.7) |
| Sex | *N male (%)* | 29/42 (69%) | 17/22 (77%) | 12/20 (60%) |
| Censored? | *N (%)* | 5/42 (12%) | 4/22 (18%) | 1/20 (5%) |
| Number of days before censoring | *Mean (CI)* | 39.4 (59.4-19.4) | 44.3 (66.2-22.3) | 20 (NA) |
| Days from symptom onset to ICU | *Mean (CI)* | 6 (8.3-3.7) | 7.5 (9.9-5.2) | 4.2 (8.5-0) |
| APACHE II on ICU admission | *mean (CI)* | 25.3 (27.6-22.9) | 23.7 (27-20.4) | 27 (30.5-23.5) |
| Mean of SOFA score for 1st 3 days | *mean (CI)* | 9.6 (10.7-8.5) | 9.3 (11-7.7) | 9.9 (11.6-8.3) |
| Mean of SOFA score for 1st 7 days | *mean (CI)* | 8.9 (10.1-7.8) | 9.1 (11-7.3) | 8.7 (10.3-7.2) |
| ICU days (censored) | *mean (CI)* | 14.1 (17.3-10.8) | 14.2 (20.5-7.8) | 14 (16.9-11.1) |
| Death in ICU | *N (%)* | 13/42 (31%) | 7/22 (32%) | 6/20 (30%) |
| Mechanical ventilation days<br>(censored) | *Mean (CI)* | 14.4 (18.9-10) | 16.8 (25.1-8.6) | 11.8 (14.9-8.7) |
| Total days of ventilation rescue<br>measures | *Mean (CI)* | 2.9 (4.3-1.4) | 4.4 (7-1.8) | 1.2 (2-0.4) |
| ANA (admission and up to 10d) | *N (%)* | 27/42 (64%) | 15/22 (68%) | 12/20 (60%) |
| ANA titre? | *N (%) 0 to 80* | 15/42 (36%) | 7/22 (32%) | 8/20 (40%) |
|  | *N (%) 160 to 320* | 16/42 (38%) | 9/22 (41%) | 7/20 (35%) |
|  | *N (%) >320* | 11/42 (26%) | 6/22 (27%) | 5/20 (25%) |
| Highest ANA titre | *mean (CI)* | 655 (888-422) | 677 (1021-334) | 629 (986-273) |
| AC4 and/or AC5 pattern | *N (%)* | 26/42 (62%) | 13/22 (59%) | 13/20 (65%) |
| AC19 and/or AC20 pattern | *N (%)* | 15/42 (36%) | 10/22 (45%) | 5/20 (25%) |
| spAAB | *N (%)* | 16/42 (38%) | 9/22 (41%) | 7/20 (35%) |
| **Myositis-related spAAB** | *N (%)* | 13/42 (31%) | 6/22 (27%) | 7/20 (35%) |
| **Ever high-positive anti-cytokine AAB?** | *N (%)* | 16/42 (38%) | 11/22 (50%) | 5/20 (25%) |
| **Ever positive anti-cytokine AAB?** | *N (%)* | 22/42 (52%) | 14/22 (64%) | 8/20 (40%) |

Abbreviations: AC, International Consensus on Autoantibody Patterns anti-cellular pattern nomenclature; ANA, anti-nuclear antibodies; APACHE, Acute Physiology and Chronic Health Evaluation (score); CI, 95% confidence interval; ICU, intensive care unit; IQR, 25—75% interquartile length; SOFA, sequential organ failure assessment; spAAB, antigen-specific autoantibodies. For more details on the nomenclature used for the autoimmune assays please refer to the online materials and methods.

The study started on March 26^th^, 2020, and the first patient was recruited on March 29^th,^ 2020. The study is ongoing; the last patient from the cohort in this manuscript was recruited on May 17^th^, 2020, and the data was censored for analysis on May 31^st^, 2020. No specific COVID-19 therapeutic interventions were administered as these patients were treated prior to the availability of evidence-based treatment guidelines demonstrating effective therapies. Patients in the study cohort were separated into COVID-19 positive (COVID^+^) and non-COVID-19 (COVID^−^) based on the RT-PCR results.

### Data and sample collection

Demographics, clinical data and clinical laboratory results were collected from the patients’ paper and electronic medical records. Blood samples were collected longitudinally starting immediately upon admission if possible (defined as day 0), and on the morning of days 1, 3, 5, 7 and 10; after day 10 or ICU discharge, they were sampled every 2 weeks. Clinical data were obtained at the same times even if blood samples were unavailable. Blood samples were collected, handled and processed in rigorous standard operating procedure in a dedicated translational research station located inside the ICU (see online materials for details). To analyze longitudinal trends, only patients with 3 or more sequential samples were included in the study. To mitigate bias, five patients with shorter ICU admissions were included; 2 had early deaths and 3 had early discharges.

### Experimental procedures

All autoantibody and serology assays were performed by Mitogen Diagnostics Laboratory (MitogenDx, Calgary, AB, Canada). A HEp-2 indirect immunofluorescence assay (IFA) was used to detect anti-cellular antibodies (referred to as anti-nuclear antibodies (ANA); NOVA Lite HEp-2, Inova Diagnostics, San Diego, CA) and images read and archived on an automated instrument (Nova View, Inova Diagnostics).

All samples were tested for systemic autoimmune disease-related AAB by a FIDIS Connective13 addressable laser bead immunoassay (TheraDiag, Paris, France) and read on a Luminex 200 system using the MLX-Booster software. Anti-dsDNA positivity and titers were detected by a chemiluminescence test (Bio Flash®, Inova Diagnostics, San Diego, USA).

All samples were also tested for AAB associated with autoimmune inflammatory myopathies using a multiplexed solid-phase line immunoassay (Euroimmun AG, Luebeck, Germany), and anti-NT5c1A by addressable laser bead immunoassay. Anti-Cytokine antibodies were assayed using a multiplexed addressable laser bead immunoassay (Millipore, Oakville, ON, Canada; HCYTAAB-17K-15) on a Luminex 200 system.

### Nomenclature

AAB is a general term that encompasses the autoimmune humoral responses assayed. The HEp-2 IFA are commonly referred as anti-nuclear antibodies (ANA) even though cytoplasmic and cell cycle patterns were included in the analysis. Their classification was according to the International Consensus on Autoantibody Patterns (ICAP) nomenclature (www.anapatterns.org: last accessed 1/29/2021). The AAB test results that identified specific, named target antigens (see details above), were called collectively specific AAB (spAAB). We have further separated them into myositis-related and non-myositis-related AAB. Anti-cytokine AAB are referred to directly.

### Data analysis

All the data was organized by UT and analyzed by UT and ADB. ANOVA was used for continuous variables and Fisher’s exact test was used for categorical variables at α=0.05, adjusted for multiple comparisons as indicated in the text using the false discovery rate at q=0.05. Bayesian analysis: a Gibbs Markov chain Monte Carlo sampling was employed to estimate posterior distributions of the difference between COVID^+^ and COVID^−^, using non-informative priors. The mean difference and 95% high density intervals (HDI) were calculated. A result was considered significant for a given variable if the 95% HDI of the difference between COVID^+^ vs. COVID^−^ patients did not enclose zero. An exploratory analysis was conducted to estimate the effect different priors would have on the posterior distributions; the priors represent varying pre-existing assumptions on the prevalence of differences between COVID^+^ vs. COVID^−^ patients. All statistical and graphical analyses were performed on R (RStudio, version 1.3.1093, Boston, United States) and JMP Pro (version 15.2.1; SAS Institute Inc, Cary, NC, USA).

### Online methods

For additional details, see the online methods.

## Results

The demographic and clinical characteristics, including past medical history are shown in Table 1 (see also Table S1). No statistically significant clinical differences were noted between the two patient groups. Importantly, both patient groups experienced comparable disease severity^11, 12^ as measured in ICU days, mechanical ventilation days and mortality rates, as well as surrogate severity scores (Table 1). Age, sex and ethnicity were not correlated with the presence of AAB (not shown).

The presence of ANA in general were not significantly associated with disease severity (Figure 1a), although there was a positive correlation with disease severity that did not reach statistical significance. Nevertheless, the presence of ICAP AC19 (cytoplasmic dense fine speckled) and/or AC20 (cytoplasmic fine speckled) IFA patterns, specifically, were consistently associated with worse severity of illness scores in both patient groups (Figure 1a). Neither the prevalence nor the IFA staining patterns were different between the COVID^+^ and COVID^−^ groups, although some patients demonstrated unique IFA patterns (Figure 1b).

**Figure 1.**
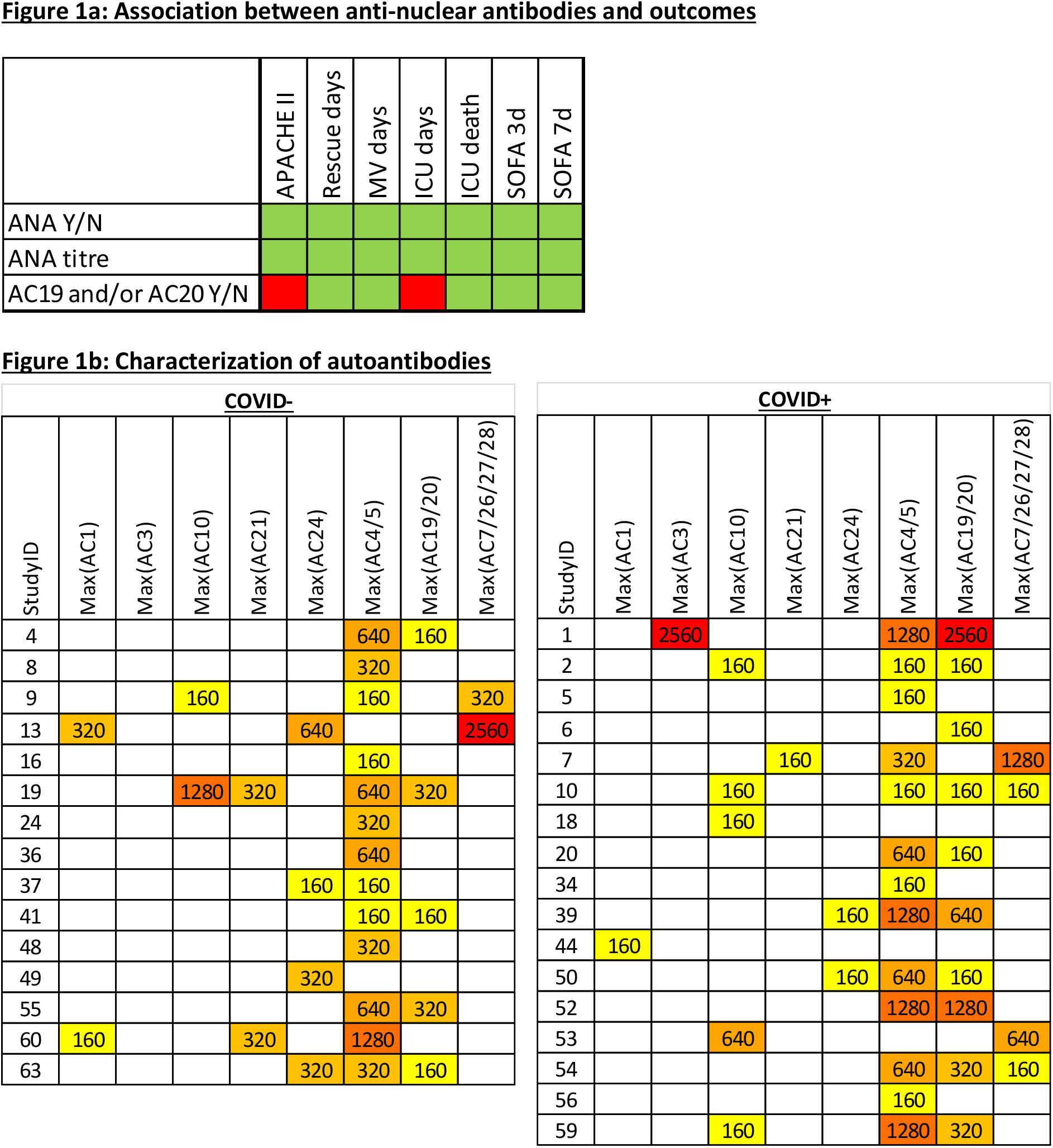
Anti-nuclear antibodies. Abbreviations: ANA, anti-nuclear antibodies; AC, International Consensus on Autoantibody Patterns anti-cellular pattern nomenclature; APACHE, Acute Physiology and Chronic Health Evaluation (score); Max, end point titer; ICU, intensive care unit; MV, mechanical ventilation; SOFA, sequential organ failure assessment (score). For more details on the nomenclature used for the autoimmune assays please refer to the online materials and methods. <u>Legend:</u> A) Association between anti-nuclear antibodies and outcomes. Refer to the legend of Table 1 for details on the variables. AC19 is shown together with AC20 given their similarity, as is the common usage. The variables were compared in a pairwise fashion using ANOVA for continuous variables and Fisher’s exact test for categorical variables at α=0.05, followed by the false discovery rate at q=0.05; green squares indicate lack of association, red squares indicate statistically significant associations. Y/N denotes dichotomization into whether they were present “yes or no” (i.e. titre above 1:80). Similar to table 1, the ANA represent the results during longitudinal sampling up to day 10 of ICU admission, for standardization among patients. The clinical outcomes were measured for up to 3 months. B) Characterization of autoantibodies. Heatmap representation of the highest ANA titers during the first 3 months of admission. Includes more patients than in Table 1 and Figure 1a since some AABs developed after 10 days. This may introduce bias since not all patients had prolonged hospitalizations but is a better representation of the dynamics and spectrum of ANA detected. Only patients with titers > 1:80 are included. AC4 is shown together with AC5, and AC19 is shown together with AC20, given their similarity, as is the common usage. AC7, 26, 27 and 28 are shown together given their rarity individually.

We compared the incidence of spAAB directed against autoantigens typically associated with systemic autoimmune diseases and myositis in critically ill COVID^+^ and COVID^−^ patients (Figure 2). Patients who had a single, transient, low-titer spAAB result were not classified as having a positive test. Figure 2 shows that the presence of spAAB was similar in both COVID^+^ and COVID^−^. In addition, there was no difference between the temporal development of spAAB between the two groups (Figure 2), nor was there a correlation between the emergence of AAB and SARS-CoV-2 seroconversion in the COVID^+^ group (not shown). Of note, most of these AAB persisted beyond 10 days (Figure 2), with the longest measurement available up to 54 days of hospitalization. There was no correlation between spAAB with COVID-19 status (Table 1) or any of the measures of disease severity.

**Figure 2.**
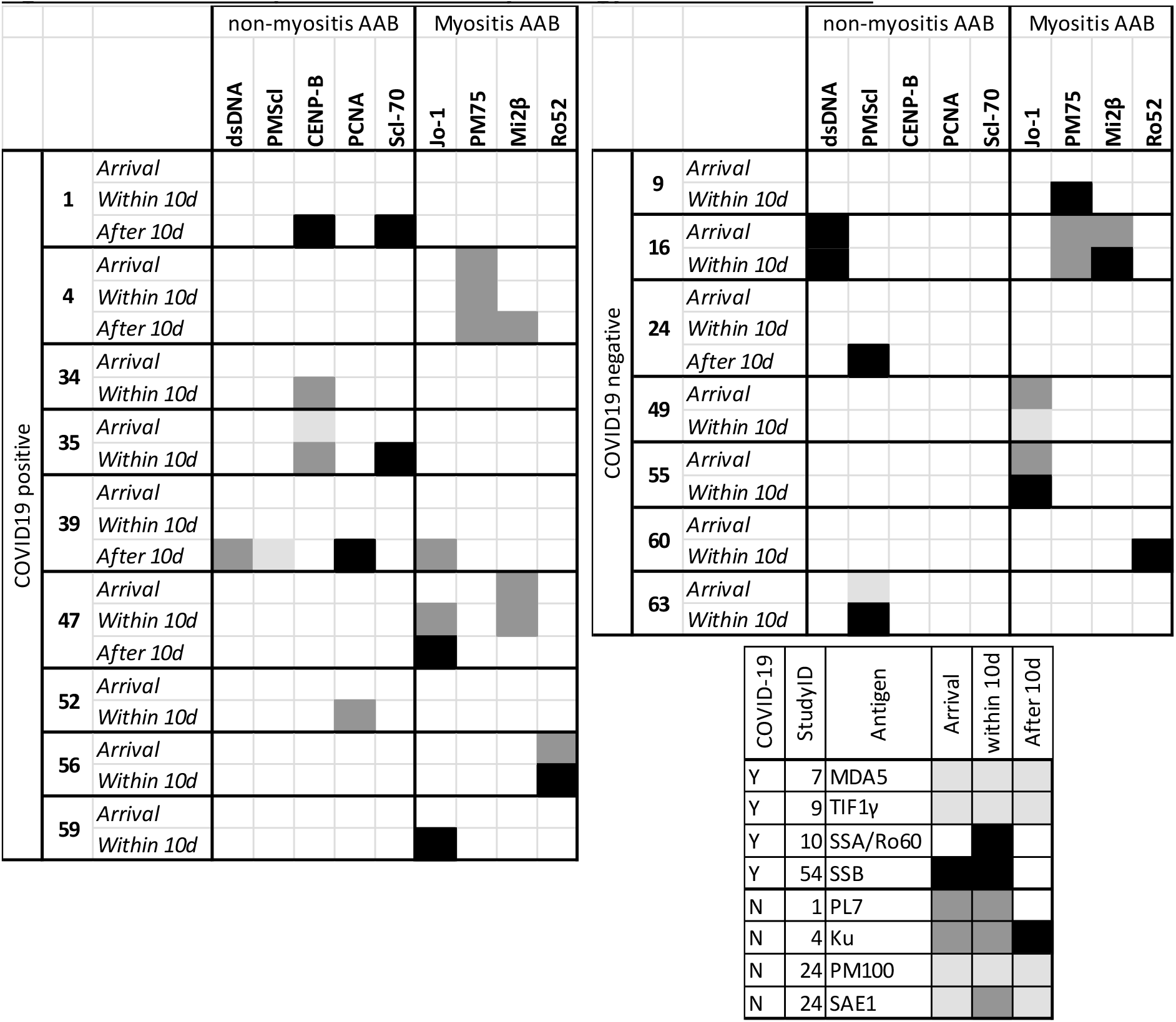
Evolution of specific autoantibody serology and titres over time. Abbreviations: AAB, autoantibodies; CENP-B, centromere protein; dsDNA, double stranded DNA; Jo-1, histidyl tRNA synthetase; Ku, a component of the DNA-dependent protein kinase complex; Mi2β, a component of the nucleosome remodeling-deacetylase complex; MDA, melanoma differentiating antigen 1; PCNA, proliferating cell nuclear antigen; PL7, threonyl-tRNA synthetase; PM75/100, polymyositis-scleroderma (PM/Scl) overlap antigens part of the nucleolar PM/Scl complex; Ro52: tri-partite motif 21 antigen; SAE1, component of the small ubiquitin like modifier activating enzyme heterodimer; Scl-70, DNA topoisomerase I; SSA/Ro60, Sjögren syndrome antigen A; SSB, Sjögren syndrome antigen B. For more details on the nomenclature used for the autoimmune assays please refer to the online materials and methods. <u>Legend:</u> The left panel shows the specific AAB serology of COVID^+^ patients, the right panel shows the specific AAB serology of COVID^−^ patients, and the lower panel shows specific AAB that were detected on only one patient, shown separately for clarity. The titres were classified into low, medium and high levels using cutoffs established by the laboratory, shown in light grey, dark grey and black, respectively; white indicates negative. Arrival indicates the first sample obtained, either on the day of admission or the next morning. Within 10 d indicates during the longitudinal sampling, up to day 10 of ICU stay. After 10 d indicates later samples, collected every two weeks; not all patients had samples after 10 days. The following specific AAB were not detected in any samples: histones, Sm/U2-U6 RNP, U1-RNP, ribosomal P, topoisomerase I, Mi2-α, MDA5, NXP2, PL7, PL12, SRP, EJ, OJ, HMGCR or NT5C1 A/Mup44 and SAR1.

50% of COVID^+^ patients had high titers of anti-cytokine AAB, as compared to 25% for COVID^−^ (Table 2). This difference was driven mainly by anti-interleukin (IL)-6, anti-IL-10 and anti-IL-17f AAB but it did not reach statistical significance (Tables 1 and 2). When analyzing all positive results of anti-cytokine AAB (as opposed to only the high titres), the difference between COVID^+^ and COVID^−^ remained at ∼25% (Table 1). Interestingly, COVID^−^ patients had a similar incidence of anti-interferon-γ and anti-interferon-β AAB as COVID^+^ (Table 2). Anti-cytokine AAB were not associated with the presence of spAAB or ANA (not shown).

**Table 2.**
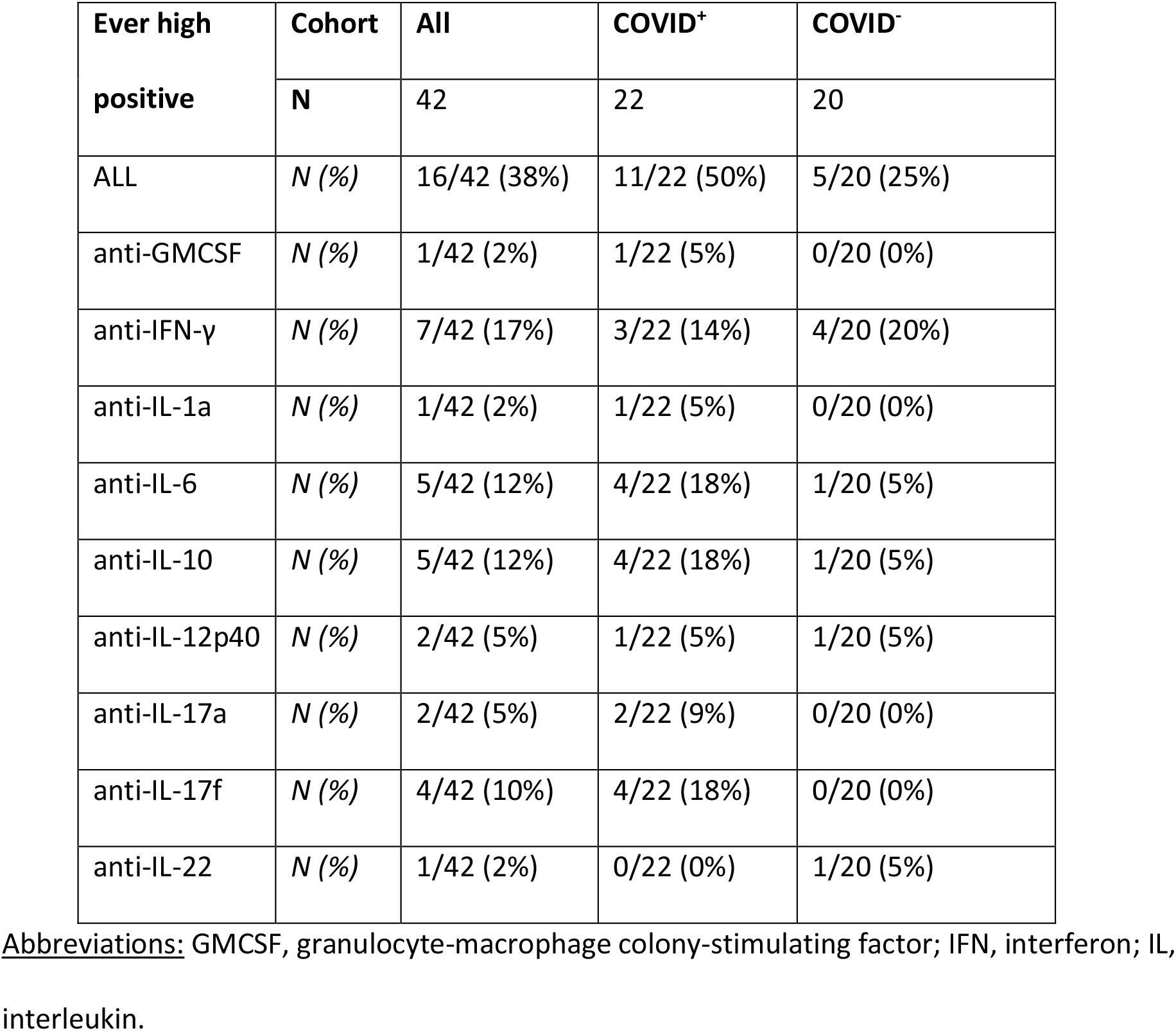
High titre anti-Cytokine autoantibodies <u>Legend:</u> The table reports the number of patients with high titres of anti-cytokine antibodies at any time during the first 10 days of ICU admission. “ALL” represents all patient having a high titre of anti-cytokine antibody of any type during that period. The numbers of the specific anti-cytokine antibodies sum to more than “ALL” since some patients had more than one high titre anti-cytokine antibody. Once adjusted for multiple comparisons, there were no statistically significant differences between COVID+ and COVID-for any of the results (Fisher’s exact test at α=0.05 followed by the false discovery rate at q=0.05). The following anti-cytokine AAB did not show high levels in any of the patients: anti-BAFF, anti-IFN-β, anti-TNF-α, anti-IL8, anti-IL-15 and anti-IL-18.

To better understand the ramifications of our data as well as confirm our results, we performed a Bayesian analysis, which is more information-rich than null hypothesis statistical testing (NHST)^13^. In Figure 3a we show the distribution of posterior probabilities for the difference between the COVID^+^ and COVID^−^ cohorts for ANA, spAAB and anti-cytokine AAB. Since all the 95% credible HDI cross the zero, our data is compatible with the null hypothesis, confirming the results from the NHST analysis. Importantly, this analysis used a non-informative prior; that is, we did not assume any previous knowledge of what the true state of nature is. This is fitting since there little is known on the prevalence of autoimmune phenomena among critically ill patients^5^ and, to our knowledge, this is the first report on autoimmunity among severe COVID-19 patients that has a control group of similarly ill, critical patients. We then performed an exploratory analysis assigning speculated, *a priori* differences between the COVID^+^ and COVID^−^ populations – Bayesian prior probabilities. Figure 3b shows that, for our results to be compatible with the existence of a significant difference, it would be necessary to assume a prior probability of the COVID^+^ patients having a prevalence that is at least 15% higher for anti-cytokine AAB, 20% higher for spAAB, and 35% higher for ANA. The resulting posterior mean differences would be in the order of 15 to 20% more autoimmunity among COVID^+^ than COVID^−^ (summarized in Table S2).

**Figure 3.**
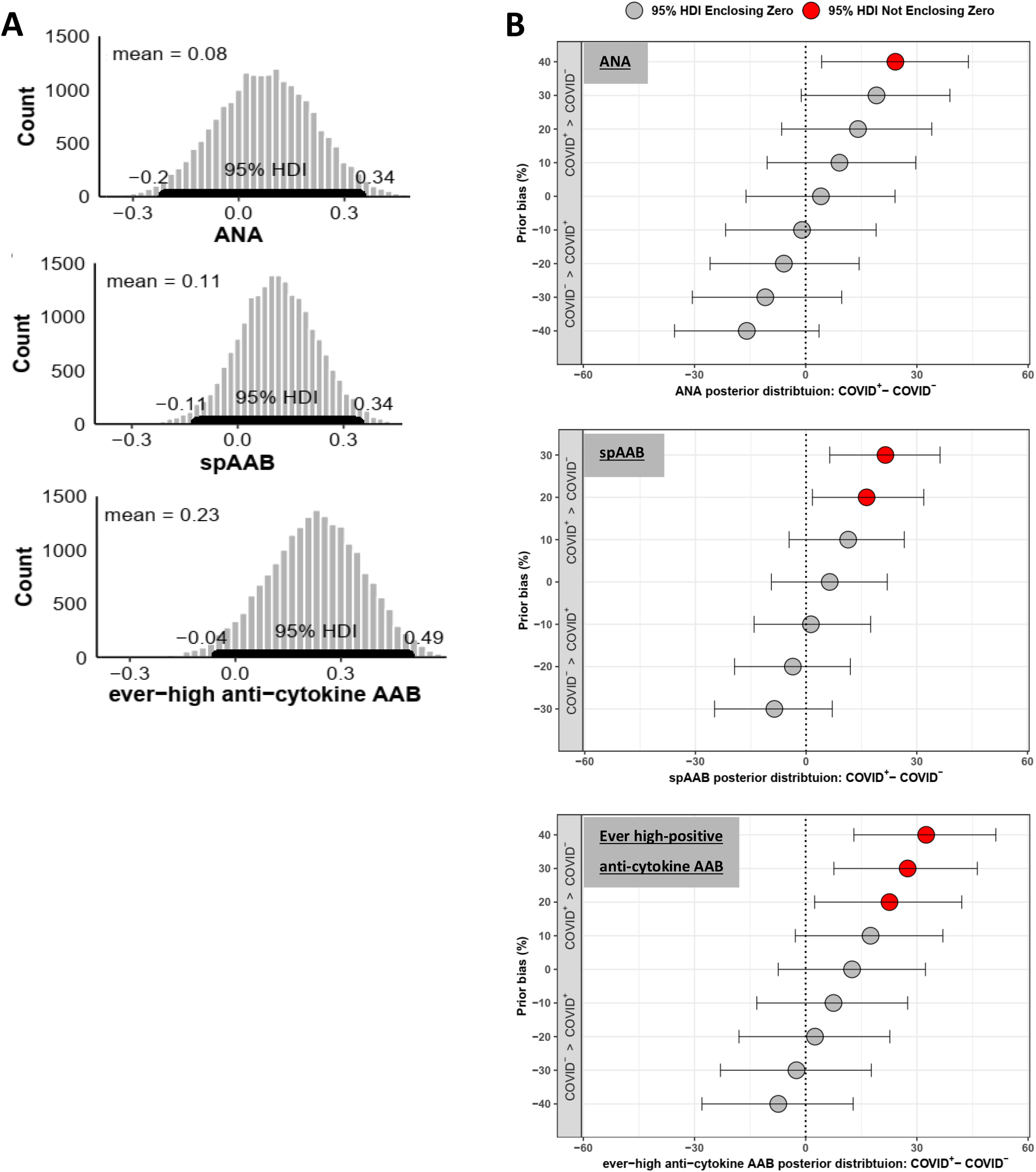
Bayesian analysis of the 95% high density credible interval of the difference between COVID^+^ and COVID^−^ levels of autoimmune serology. Abbreviations: AAB, autoantibody; ANA, anti-nuclear antibodies; AAB, autoantibodies; HDI, high density interval; spAAB, antigen-specific autoantibodies. <u>Legend:</u> A) Posterior distributions of the difference in humoral autoimmunity in COVID^+^ vs. COVD^−^ patients. The histograms plot the difference between the COVID^+^ and COVID^−^ in the proportion of patients who are positive for the variables presented. The 95%HDI is visually represented by the bold black line along the x-axis. The histograms represent simulations using four Gibbs sampling chains employing a flat prior probability, at 5000 iterations per chain, minus 1% burn-in, resulting in 19800 iterations in total for each histogram. B) Effect of prior bias on posterior distributions. The data was generated using the same procedure as in “A”, with the X axis representing the difference in the posterior distribution between the COVID^+^ and COVID^−^. The flat prior probability was exchanged varying levels of prior bias; the box and whiskers represent the 95% HDI at each level of prior bias. The positive Y axes show the results assuming that COVID^+^ have a higher incidence of positivity than COVID^−^, while the negative Y axes show the opposite; the zero Y indicates a prior that assumes no bias, with an equal probability of 50% positivity rate for both COVID^+^ and COVID^−^. A 95% HDI that does not include the zero on the X axis indicates a significant difference between the COVID^+^ and COVID^−^ posterior probability distributions at that level of prior bias, indicated with a red dot. The simulations were performed on 5% intervals; the results are shown for 10% intervals for clarity.

## Discussion

We analyzed disease severity and presence of AAB in 22 COVID^+^ and 20 COVID^−^ ICU patients with respiratory distress of similar severity, enrolled contemporaneously, over time. We used HEp2 IFA to typify ANAs, as well as solid-phase multi-analyte arrays and bead-based multiplexed immunoassays to typify specific autoreactive antigens such as those observed in autoimmune diseases such as systemic lupus erythematous, Sjögren syndrome, systemic sclerosis, autoimmune inflammatory myopathies, and others^4-6^.

Overall, close to 15,000 data points were collected between demographic variables and laboratory and clinical variables over time. We observed high prevalence and titres of AAB among the COVID^+^ patients our study (68% had ANA), in agreement with some previous reports of COVID^+14, 15^. Yet, surprisingly, we also observed high prevalence and titres of AAB in the COVID^−^ patient cohort (60% had ANA). In another cross-sectional study^4^, the majority of positive sera had reactivity to only single nuclear antigens, whereas most of our patients showed multiple reactivities (Figure 2), suggesting a relatively widespread loss of humoral tolerance in some of our patients. We found no major differences in either the autoantigen specificity, temporal dynamics, or titers in COVID^+^ vs. COVID^−^ cohorts. These data suggest that AAB production may be a feature of immune dysfunction associated with acute systemic illness rather than specifically with a SARS-CoV-2 driven immune pathology. Importantly, specific ANA patterns (AC19 and/or AC20) were broadly correlated with disease severity measures, suggesting a causal relationship, or at least a common factor “driving” autoreactive B cell responses in critically ill patients.

A previous study reported that approximately 10% of patients with life-threatening COVID-19 pneumonia had neutralizing IgG AAB against a spectrum of interferon proteins, which blocked SARS-CoV-2 infection *in vitro*^16^. Of note, these AAB were not found in asymptomatic or mild SARS-CoV-2 infection, but they were not studied in similarly ill, COVID-patients. Our results show that matched COVID^−^ controls have a similar incidence of anti-interferon-γ and anti-interferon-β AAB as COVID^+^ patients. While we did not study if the anti-cytokine AAB in our study had neutralizing activity, we did not find that the presence of these AAB differed between COVID^+^ and COVID^−^. The small sample size and the large absolute difference for anti-IL-6, anti-IL-10 and anti-IL-17f raise the possibility of a type-2 error. We addressed this concern using Bayesian analysis, which confirmed the results of the NHST.

Our data and analysis support that, humoral autoimmunity, as described in detail using an extensive panel of ANA, spAAB and anti-cytokine AAB, longitudinally, does not seem to be a particular characteristic of COVID+ patients but rather of critically ill patients with respiratory failure. The collection of data over time significantly increases the reliability of our results by a) reducing the biases of different, arbitrary sampling times between different patients, b) providing test-retest internal validity, and c) characterizing the development of autoimmunity in a physiological context - over time. Although there is no direct evidence to assume *a priori* that COVID^+^ have more autoimmunity than COVID^−^, some clinicians may consider the immune characteristics of severe COVID-19 enough reason to warrant such assumptions. Our analysis suggests that, in that case, the expected differences would be in the order of a modest 15-20% (Figure 3b, Table S2). These results provide boundaries to the autoimmunity phenomenon of COVID^+^ patients and provide the context of high prevalence of autoimmunity among critically ill patients with respiratory failure in general. Nevertheless, there are some suggestions in our data that, within this context, COVID-19 patients may have a tendency for particular autoimmune phenotypes, as hinted by the large absolute difference of anti-IL6, anti-IL10 and anti-IL17f AAB. Larger studies are needed to assess this possibility, as well as the pathogenic role of AAB in both COVID^+^ and COVID^−^ patients, which is yet to be demonstrated.

Given that all of our patients (both COVID^+^ and COVID^−^) had respiratory failure, it may be that the association between AAB production is related to lung injury and/or lung endothelial pathology, a known feature of acute lung injury^17^ regardless of etiology. The activation of both innate immune and adaptive responses to abnormal danger-associated molecular patterns released into the circulation following the initial lung injury may contribute to the loss of humoral tolerance^18-21^. Another potential cause of AAB formation may be related to immune activation associated with the biotrauma of ventilator associated lung injury, in the context of loss of pulmonary compartmentalization^22^. This could explain some of the AAB that developed during the course of hospitalization (figure 2). Indeed, perhaps the most salient result of our study is the high prevalence of broad autoimmune serology among the critically ill, a phenomenon we describe for the first time here. These results stand in stark contrast to the known prevalence in the general population of about 5%, bearing further study.

Our study’s main shortcoming is the small sample size. We account for this by employing Bayesian analysis to explore the boundaries of the possible states of nature underlying our results. Another shortcoming is the need to consolidate the different autoimmune serologies under the broad categories of ANA, spAAB and anti-cytokine AAB, due to the small sample size and to avoid multiple testing. Nonetheless, we do present these results in detail, providing a qualitative assessment, and performed analyses of specific features when the sample size allowed for it, such as for the AC19 and/or AC20 patterns, or the ANA titres.

In conclusion, in some patients, severe COVID-19 is characterized by immune dysregulation with clinical features of systemic autoimmunity. We present evidence that humoral autoimmunity is a common feature of critically ill patients with respiratory deterioration. Our study urges caution in the interpretation of AAB test results among COVID^+^ patients. Further studies are needed to determine whether transient or sustained production of AAB in the setting of acute illness is associated with the development of systemic autoimmunity.

## Data Availability

The datasets used and/or analysed during the current study are available from the corresponding author on reasonable request.

## Declarations

### Ethics approval and consent to participate

- This research was approved by Research Ethics Boards at St Michael’s Hospital and performed in accordance with the Helsinki Declaration of 1975 as revised in 2013.

### Availability of data and materials

- The datasets used and/or analysed during the current study are available from the corresponding author on reasonable request.

### Authors’ contributions

- This report is part of the COLOBILI study (Coronavirus longitudinal biomarkers in lung injury). AB and CDS are principal investigators; MJF, RR and AS are collaborators/co-investigators and UT is the research lead.
- RR, MJF, UT, and CDS conceived of the study; MJF, UT and RR wrote the manuscript drafts; AS, AB and CDS provided critique and technical guidance; UT and ADB performed the data analysis and creation of the figures. All authors edited the manuscript, through to the final version, read and approved the final submission.

## Acknowledgements

- The authors acknowledge the technical assistance of Haiyan Hou, Meifeng Zhang and Emily Walker in the MitogenDx Laboratory at the University of Calgary. We thank Marlene Santos, Gyan Sadhu, Imrana Khalid, and Sebastian Duncan, the research coordinators at St Michael’s Hospital Critical Care Research Unit. We are grateful to patients and families that have generously consented to the study.

## Study registration

- NCT04747782 – clinicaltrials.gov

