## Supplementary material for "COVID-19 associated autoimmunity is a feature of severe respiratory disease - a Bayesian analysis": Suplemental Materials and Methods and Supplemental Tables 1 and 2

**<u>Table of Contents:</u>**

1) Online Materials and methods – page 2
2) Supplemental table 1 – page 8
3) Supplemental table 2 – page 9

### Online materials and methods

**Study design.** This report is part of the COLOBILI study – Coronavirus Longitudinal Biomarkers in Lung Injury, being conducted at St. Michael's Hospital (Toronto, ON, Canada). This is an observational cohort study that includes analysis of biological samples. The study was approved by the Research Ethics Board of St. Michael's Hospital (REB# 20-078). The inclusion criteria were all patients above age 18 years admitted to the Medical-Surgical or Trauma-Neuro intensive care units (ICU) with acute respiratory distress, suspected to have COVID-19. COVID-19 status was determined according to diagnostic PCR of nasopharyngeal swabs and/or endotracheal aspirates as described in detail below. The exclusion criteria were refusal to participate, inability to ascertain mortality status during the first 2 weeks of the study, failure to obtain a blood sample on either day 0 or 1, or individuals known to have had COVID-19 in the 4 weeks prior to admission in any setting. Patients were followed for up to 3 months in hospital or hospital discharge, whichever occurred first. The primary outcome was death in the ICU; secondary outcomes included death outside the ICU, ICU utilization metrics, and organ dysfunction measures and scores. Clinical data and blood samples were collected longitudinally immediately upon admission, as available, defined as day 0, and on the morning of days 1, 3, 5, 7 and 10; after day 10 or ICU discharge, they were sampled every 2 weeks. The study started on March 26th, 2020, and the first patient was recruited on March 29th, 2020. The study is ongoing; the last patient from the cohort presented in this manuscript was recruited on May 17th, 2020, and the data was censored for analysis on May 31st, 2020. No COVID-19 treatments were given to the patients beyond the standard of care since at the time there was no evidence of efficacy for any such treatments. Informed consent was obtained from the patients or their legal representatives; in case that was not possible, the patients were enrolled using a deferred consent model and kept in the study until they regained capacity, or a surrogate decision maker was identified.

**Data and sample collection.** Demographics, clinical data and clinical laboratory were collected from the patients' paper and electronic medical records, with auditing performed reciprocally by research coordination team members and curated by UT. To standardize handling and processing, blood samples were collected in EDTA tubes between 8:00 and 12:00 AM and kept on ice for up to 60 minutes until their processing in a dedicated translational research station located inside the ICU. They were then immediately frozen at -20 $^{0}$C on site, and transferred to -80 $^{0}$C for storage within 48 hrs. All procedures were performed by dedicated research personnel. Nasopharyngeal samples were obtained from all patients by bedside nurses and analyzed by the clinical laboratory using either the Altona RealStar SARS-CoV-2 RT-PCR Kit 1.0 or Cepheid GeneXpert Xpert Xpress SARS-CoV-2 assay. Endotracheal tube aspirates were analyzed using the Seegene Allplex 2019-CoV Assay. All patients had a nasopharyngeal PCR performed; intubated patients had an endotracheal aspirate sent as well. Further PCR tests were repeated by the clinical and infection control teams at their discretion if there was suspicion of a false negative result based on clinical observations or to confirm negativity. All patients in the PCR negative cohort had at least two negative tests performed acutely, except one patient who had only one test done acutely. In order to analyze longitudinal trends, only patients with 3 or more longitudinal sampling times were included in the study. To mitigate bias, five patients with shorter ICU admissions were included; 2 had early deaths and 3 had early discharges.

**Experimental procedures.** Plasma samples were stored and managed under a standard operating procedure which included shipping on dry ice and storage at -80C until assay performance by Mitogen Diagnostics Laboratory (MitogenDx, Calgary, AB, Canada). A HEp-2 indirect immunofluorescence assay (IFA) was used to detect anti-cellular antibodies (also referred to as anti-nuclear antibodies (ANA) – see "nomenclature" below) [1] (NOVA Lite HEp-2, Inova Diagnostics, San Diego, CA) at a serum dilution of 1:80 and read on an automated instrument (Nova View, Inova Diagnostics) which interpolates fluorescence intensity to an end point titer[2]. IFA staining patterns were classified according to the International

Consensus on Autoantibody Patterns (ICAP, https://anapatterns.org/index.php)[3]. All samples were also tested for systemic autoimmune disease-related autoantibodies by a FIDIS Connective13 addressable laser bead immunoassay (ALBIA, TheraDiag, Paris, France) detecting antibodies to Sm/U2-U6 ribonucleoprotein (RNP), U1-RNP, SSA/Ro60, SSB/La, Ro52/Tripartite Motif Protein 21 (TRIM21), histones, and ribosomal P, read on a Luminex 200 system using the MLX-Booster software. A cut-off of >40 units was considered positive. Anti-dsDNA positivity and titers were detected by a chemiluminescence test (Inova Diagnostics, San Diego, USA). A cut-off of <27 chemiluminescence units was considered within normal range, 27-35 was indeterminate, and >35 was positive. All samples were also tested for autoantibodies associated with autoimmune inflammatory myopathies using a multiplexed solid phase line immunoassay: Ro-52/TRIM21, OJ, EJ, PL-12, PL-7, SRP, Jo-1, PM-75, PM-100, Ku, SAE1, NXP2, MDA5, TIF1γ, Mi-2α, Mi-2β (Euroimmun AG, Luebeck, Germany), and anti-NT5c1A by addressable laser bead immunoassay[4]. The following anti-cytokine antibodies were assayed using an addressable laser bead immunoassay (Millipore, Oakville, ON, Canada; HCYTAAB-17K-15) read on a Luminex 200 system: BAFF, GMCSF, IFN-β, IFN-γ, IL-1a, IL-6, IL-8, IL-10, IL-12p40, IL-15, IL-17a, IL-17f, IL-18, IL-22 and TNF-α. The manufacturer's thresholds were 500 for positive and 1000 for high-positive (arbitrary units). All tests were performed according to the manufacturer's instructions.

**Nomenclature.** There is considerable heterogeneity in the nomenclature of autoimmune assays in the literature and clinical practice; therefore, we used the most contemporary usages. Autoantibodies is a general term that encompasses all of the autoimmune humoral responses assayed. The HEp-2 IFA, although including anti-cytoplasmic and anti-mitotic cell antibodies, are commonly referred as anti-nuclear antibodies (ANA), and we have adopted that usage for clarity. Their classification uses the different patterns identified as AC0, AC1, AC2, etc. according to the International Consensus on Autoantibody Patterns (ICAP: https://anapatterns.org/index.php)[5]. The AAB test results that identified specific, named antigens (see details above), were called collectively specific autoantibodies (spAAB).

We have further separated them into myositis-related and non-myositis-related AAB. Anti-cytokine antibodies are referred to directly.

**Data analysis.** All the data was organized by UT and analyzed by UT and ADB. The data was censored on May 31st, 2020; only 5 patients had censored data for the primary outcome, death in the ICU within 3 months. Given the elapsed time until censoring, the risk of right-censoring bias is low. ANOVA was used for continuous variables and Fisher's exact test was used for categorical variables at $\alpha$=0.05, adjusted for multiple comparisons as indicated in the text using the false discovery rate at q=0.05. Bayesian analysis: To evaluate the difference in the proportion of individuals who were positive for AAB in COVID$^+$ vs. COVID$^-$ patients a Gibbs Markov chain Monte Carlo sampling algorithm was employed. For each variable, posterior parameter estimation in both groups was derived via the Bernoulli distribution function, incorporating a uniform beta prior ($\alpha = 1$, $\beta = 1$). The simulations were run across 4 chains, each with 5000 iterations and a 1% burn-in (50 iterations removed per chain); due to the inherent acceptance of all proposed values in the Gibbs algorithm, the effective sample size for all chains was 19800. Chain convergence was measured using the Gelman convergence statistic, with the accepted convergence value of all chains set at <1.01 [6]. Monte Carlo standard error was also calculated on all simulations to measure chain accuracy [7]. Briefly, for each iteration of the simulation, a credible parameter value was obtained for both groups: the parameters ($\theta$) represented the probability of occurrence (success) for each variable, which in the current case is represented by AAB positivity. A posterior distribution of difference scores was also calculated ($\theta 2$ - $\theta 1$). From this posterior distribution, the mean difference value and 95% High Density Interval (HDI) were derived; the HDI was calculated according to the shortest credible interval method to account for potentially slight asymmetries in the distribution [8] Posterior predictive checks were employed; simulated data derived from the model was qualitatively evaluated against the observed proportions to evaluate fit. A result was considered significant for a given variable if the 95% HDI of the difference between COVID$^+$ vs. COVID$^-$ patients did not enclose zero.

Finally, an exploratory analysis was conducted to estimate the effect different priors would have on the posterior distribution of ANA, spABB and ever high-positive anti-cytokine AAB; the priors represent varying pre-existing assumptions of the prevalence differences between $COVID^+$ vs. $COVID^-$ patients. Starting at 0% difference, beta priors were employed to adjust the difference in positivity between groups. A $\kappa$ level equivalent to the sample size of each group ($COVID^+$, $\kappa = 22$; $COVID^-$, $\kappa = 20$), was used to allocate equal weight of the prior to the observed data. This avoids the introduction of arbitrary assumptions by conserving the structure of the observed data. The initial starting point was derived from the combined prevalence of each variable's positivity in all participants (n = 42, Table 1). Prior bias was then allocated symmetrically. For example, the combined proportion of all patients who were ANA positive was 64%; a 10% difference skewed towards the $COVID^+$ group was evaluated by adding 5% to $COVID^+$ and subtracting 5% from COVID- patients, resulting in a 69% vs 59% assumed prior prevalence of ANA. This was done in 5% increments until significant posterior distributions were found for all variables studied, at 40%. All statistical and graphical analyses were performed on R (RStudio, version 1.3.1093, Boston, United States) and JMP Pro (version 15.2.1; SAS Institute Inc, Cary, NC, USA).

**Table S1: Premorbid Clinical Characteristics and Therapeutics**

| | All (N) | COVID+ (N) | COVID- (N) |
|---|---|---|---|
| | 42 | 22 | 20 |
| Respiratory PMH | 18 | 8 | 10 |
| Cardiovascular PMH | 19 | 11 | 8 |
| Renal PMH | 7 | 6 | 1 |
| Type 2 Diabetes | 20 | 12 | 8 |
| Hypertension | 24 | 14 | 10 |
| Other comorbidities | 37 | 18 | 19 |
| Premorbid steroid used | 3 | 1 | 2 |
| Premorbid immunomodulatory medication use | 2 | 1 | 1 |
| Premorbid ACEi/ARB use | 15 | 10 | 5 |

Abbreviations: ACEi, Angiotensin-converting enzyme inhibitors; ARB, angiotensin receptor blocker; PMH, past medical history.

Legend: "Other comorbidities" include autoimmune diseases: Myasthenia gravis among COVID+, and autoimmune hemolytic anemia, rheumatoid arthritis and multiple sclerosis among the COVID-. No statistically significant difference between COVID+ and COVID- patients for all variables were detected using ANOVA for continuous variables and Fisher's exact test for categorical variables at $\alpha=0.05$

**Table S2: Smallest prior bias for COVID$^+$ > COVID$^-$ producing a posterior distribution with a 95% high density interval that does not include zero**

| Autoantibody | Prior bias COVID$^+$ vs COVID$^-$ | Mean Difference | 95% HDI (range, %) |
|---|---|---|---|
| ANA | 35% (76.5 vs. 51.5) | 21.5% | 1.9 – 41.2 |
| spAAB | 20% (27.0 vs 7.0) | 16.4% | 1.7 – 31.9 |
| Ever high-positive anti-cytokine AAB | 15% (45.5 vs 30.5) | 20.0% | 0.1 – 40.2 |

Abbreviations: AAB, autoantibodies; ANA, anti-nuclear antibodies; HDI, high density interval; spAAB, antigen-specific autoantibodies.

Legend: Exploratory analysis was performed with of varying levels of prior bias, assuming COVID+ patients have a higher incidence of autoimmune serology than COVID- patients. The bias was increased in 5% increments. The 2$^{nd}$ column shows the threshold level where the 95% HDI of the posterior distribution of COVID$^+$ - COVID$^-$ does not include the zero, with the exact biases used in parenthesis. The 3$^{rd}$ column shows the mean difference between the COVID$^+$ and the COVID$^-$ for that distribution, and the 4$^{th}$ column shows the range of the 95% HDI obtained.
